# Drug repurposing: Hydroxyurea therapy improves the transfusion-free interval in HbE/beta-thalassemia–major patients with Xmn1 polymorphism

**DOI:** 10.1101/2021.02.28.21251843

**Authors:** Debojoyti Ghosh, Amrita Panja, Dipankar Saha, Uma Banerjee, Asok Kumar Dutta, Anupam Basu

## Abstract

**Aims:** HbE/β-thalassemia is the prevalent form of severe β-thalassemia in Asian countries. Hydroxyurea (HU) is the most common drug used for the management of sickle-cell anemia but not thalassemia. Here, we aimed to assess clinical HU response among patients with HbE/β-thalassemia with respect to *Xmn1* γ^G^globin polymorphism and elucidate the association between this polymorphism and HU response efficacy.

**Methods:** We enrolled 49 transfusion-dependent patients with HbE/β-thalassemia. Fetal hemoglobin level was measured using High-performance liquid chromatography (HPLC) and complete blood count was determined pre- and post-HU therapy. Polymerase chain reaction–Restriction fragment length polymorphism (PCR-RFLP) was performed for genotyping *Xmn1* γ^G^globin polymorphism.

**Results:** A total of 30 (61.22%) patients were found to be responders, whereas the remaining 19 (38.78%) were non-responders. We found 33 patients with heterozygous (C/T) and three with homozygous mutant (T/T) genotype status. We obtained a statistically significant correlation (*p* < 0.001) between *Xmn1* polymorphism and transfusion-free interval. Patients with *Xmn1* polymorphism were found to be good responders for HU therapy and showed increased hemoglobin levels.

**Conclusions:** Our findings indicate that HU is a potential drug candidate for thalassemia management, particularly HbE/β-thalassemia. The results hold implications in repurposing HU as an effective and efficient therapy for HbE/β-thalassemia.

## Introduction

HbE/β-thalassemia constitutes the majority of all severe β-thalassemia cases worldwide (Hossain et al., 2017), with the highest frequencies observed in India, Bangladesh, Indonesia, Malaysia, Southern China, and Thailand (Noor et al., 2020). Further, it is prevalent in north-eastern states (3- >50%) and in eastern India (3– 4%) (Mohanty et al., 2013; Panja et al., 2017; Algiraigri and Kassam, 2017). HbE/β-thalassemia results from co-inheritance of the mutant β-thalassemia allele from one parent and HbE (CD26, G>A; GAC→AAG, Glu→Lys) from the other. The phenotype ranges from an asymptomatic carrier state to severe anemic conditions that require blood transfusion from the early age of life (Colah et al., 2017). However, many patients are not transfused or under-transfused due to lack of blood donors, insufficient national blood policies, and fragmented blood services and thus have poor quality of life. Yet, blood transfusion itself has major health hazards, such as iron overload, organ damage, transfusion-transmitted infections (TTIs), and antibody generation (alloimmunization) (Shah et al., 2019). Although iron overload can be prevented via iron chelation, this procedure requires a substantial financial support. Additionally, thalassemia patients with TTIs, resulting from repeated blood transfusion, have been shown to have 3.4 times high morbidity risk (*P* = 0.031). However, patients receiving adequate care for > 4 years have > 66% less mortality risk (*P* < 0.0001) (Dhanya et al., 2020). Therefore, any alternative to regular blood transfusion can be instrumental in improving the survival rate (Rujito et al., 2016).

Presence of multiple variations and clinical instability necessitates carefully customized treatment regimen for each patient. Alternative therapeutic approaches have been developed over time. Although hydroxyurea (HU) is a scheduled drug for the management of sickle cell disease (Yahouédéhou et al., 2018; Tshilolo et al., 2019), it is not routinely prescribed for all types of thalassemia (Colombatti et al., 2018) presumably because HU may not improve the management, and the response may be different among such patients (Italia et al., 2016; Iqbal et al., 2018).

The role of β-thalassemia mutations, including *α-globin* deletions, in HU response has been reported (Chatterjee et al., 2018; Biswas et al., 2019). However, there are only a few reports regarding the effect of genotype on the HU response in HbE/β-thalassemia patients in India (Italia et al., 2010; Kumar et al., 2014; Italia et al., 2019). Moreover, the records for eastern India are insufficient. Here, we aimed to elucidate the correlation between HU therapy response and *Xmn1* polymorphism among patients with HbE/β-thalassemia within the Bengali population. Furthermore, we also aimed to correlate the clinical severity parameters of HU-administered patients with *Xmn1* polymorphism.

## Materials and Methods

### Subject selection

This study was approved by the Institutional Ethics Committee of Burdwan Medical College and conducted in compliance with the Declaration of Helsinki. Written informed consent and assent forms were collected from the parents/guardians of all the participants prior to enlistment in the study. The inclusion criteria were (1) HbE/β-thalassemia subjects with monthly blood transfusion history and splenomegaly, and (2) subjects within the age of 6–12 years. Patients with complications, such as hepatic or renal diseases, or who were undergoing cancer therapy, were excluded. Consequently, 49 patients with HbE/β-thalassemia were investigated and recruited during January– December, 2014 from the Department of Paediatric Medicine, and Thalassemia Centre, Burdwan Medical College and Hospital, West Bengal, India

### Hematological investigation

The complete blood counts of the enrolled patients were determined using an automated cell counter (Sysmex K4500). Fetal hemoglobin (HbF) was quantitated via high-performance liquid chromatography by using BioRad Variant II program (Bio-Rad Laboratories, Hercules, CA, USA) before the initiation of HU administration and also periodically after a few months of administration to assess the effect of the therapy.

### HBB genotyping using Sanger sequencing

Genomic DNA was extracted from peripheral-blood lymphocytes via standard phenol-chloroform extraction. The genotypes of the enrolled patients were confirmed using Sanger sequencing (Panja et al, 2017). The resultant chromatograms were analyzed using Chromas Lite (version 2.1) software to screen for mutations.

### Genotyping of Xmn1 γ^G^globin polymorphism

PCR-RFLP was used for detecting γ^G^globin-158 C/T polymorphism. The following primers were used: 5⍰-AAC TGT TGC TTT ATA GGA TTT T-3⍰ (forward) and 5⍰-AGG AGC TTA TTG ATA ACT CAG AC-3⍰ (reverse). RFLP was performed by digesting the PCR products with *Xmn1* (ER 1531, 500 U, Thermo Fisher Scientific) at 37°C for 2 h, followed by 3%–agarose-gel electrophoresis. The gel was stained with ethidium bromide and visualized using a UV-gel documentation system.

### HU-therapy regimen

Initially, 500 mg HU (HYDREA, Bristol-Myers Squibb Laboratories, Princeton, NJ, USA) was administered daily to participants aged > 10 years and twice a week to participants aged < 10 years. They were monitored clinically and para-clinically (complete blood count, blood urea level, and creatinine level were measured every three months).

### HU response analysis

Response to HU administration was considered significant if patients who had previously required regular transfusions could maintain a hemoglobin level of > 7 g/dl and perform routine daily activities transfusion-free for > 8–12 months. The ones who required regular blood transfusion even after HU therapy were considered non-responders.

### Statistical analysis

SPSS software (v 27) for Windows was used for statistical analysis (SPSS Inc., Chicago, IL, USA). A *p*-value of < 0.05 was considered statistically significant. Student’s paired *t*-test was performed to compare numerical variables between a pair of groups.

## Results

According to the age of onset, the study participants were classified into three groups; group-1 included those who had their first blood transfusion and clinical presentation at the age of 6 months–1 year; group-2 at 1–2 years; and group-3 at > 2 years. Forty subjects underwent blood transfusion for > 2 years, of whom eight had to have transfusion at the age of 1–2 years. In the present study, the initial (pre-HU therapy) hemoglobin level was 6.1–8 g/dl in 31 participants; 4.1– 6.0 g/dl in 18. Furthermore, 34 had initial HbF levels of 0.1–20.0%; 13 had 20.1–40.0%, and the remaining two had 40.1-80% (Table 1).

**Table 1.** Different clinical parameters of the studied subjects with *Xmn1* polymorphism before starting HU treatment

| Clinical Parameters | Range | *Xmn1* Polymorphism |  |  |
| --- | --- | --- | --- | --- |
|  |  | C/C | C/T | T/T |
| Age of first onset | 6 months–1year | 1 | 0 | 0 |
|  | 1–2 years | 3 | 5 | 0 |
|  | > 2 years | 9 | 28 | 3 |
| Hemoglobin (g/dl) | 4.1–6 | 7 | 10 | 1 |
|  | 6.1–8 | 6 | 23 | 2 |
| Fetal hemoglobin (%) | 60.1–80 | 0 | 1 | 0 |
|  | 40.1–60 | 0 | 0 | 1 |
|  | 20.1–40 | 2 | 11 | 0 |
|  | 0.1–20 | 11 | 21 | 2 |

Out of the 33 with C/T polymorphism only 23 (69.7%) had a hemoglobin level of 6.1–8.0 g/dl whereas the seven with wildtype (C/C) genotype had 4.1–6.0 g/dl. (Table 1). Among the 26 subjects with heterozygous (C/T) genotype, the mean initial hemoglobin level was 6.22 g/dl. After HU therapy, the mean hemoglobin level increased to 7.03 g/dl (*p* < 0.0001). We found three subjects with homozygous mutant (T/T) genotype; their mean hemoglobin levels pre- and post-HU therapies were 6.66 and 7.76 g/dl, respectively (*p* = 0.1276). Conversely, 13 subjects who were wildtype (C/C) for *Xmn1* genotype had hemoglobin levels of 5.85 and 6.30 g/dl pre- and post-HU therapy, respectively (*p* = 0.0313) (Fig 1).

**Fig 1.**
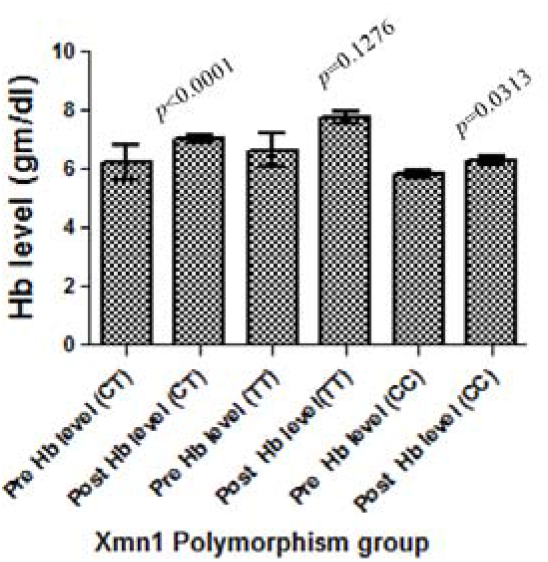
Bar diagrams showing the correlation between pre- and post-hemoglobin levels after HU treatment with *Xmn1* polymorphism among the studied Bengali HbE/β-thalassemia patients

Among the 33 subjects heterozygous for *Xmn1* polymorphism, 23 experienced >12 months of transfusion-free interval whereas three and seven underwent blood transfusion within intervals of 8–12 and 1–8 months, respectively. The three subjects with homozygous mutant (T/T) genotype underwent blood transfusion with an interval of > 12 months. The average transfusion interval was 1–4 months in most of the subjects with (C/C) genotype (11 out of 13). Therefore, our findings indicate that heterozygous (C/T) and homozygous (T/T) genotypes achieved high transfusion-free survival after HU administration. The correlation between *Xmn1* polymorphism and transfusion-free interval was statistically significant (*p* < 0.001) (Table 2).

**Table 2.**
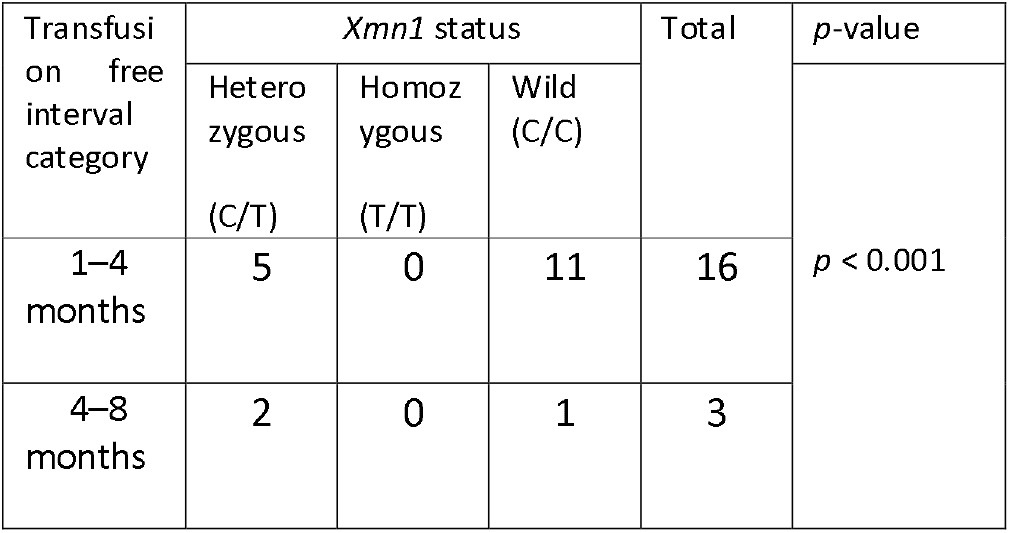
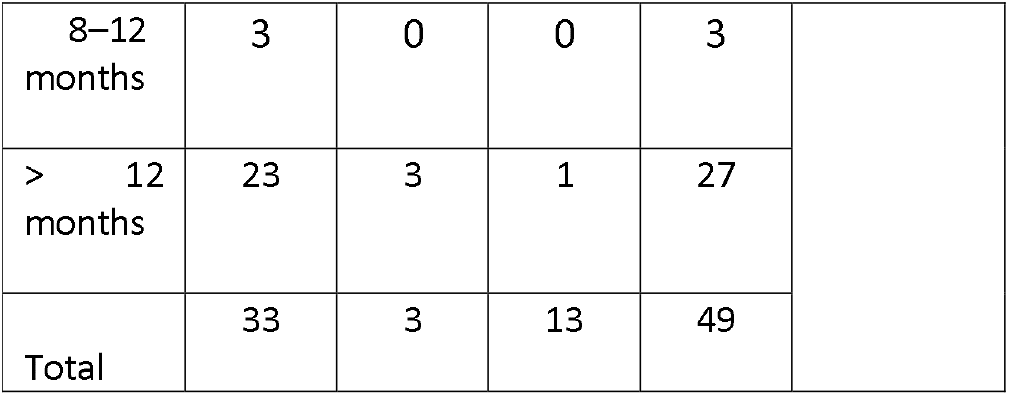
Distribution of studied subjects according to transfusion free interval category and *Xmn1 s*tatus after Hydroxyurea treatment

In this study, 30 (61.2%) patients responded to HU therapy unlike the remaining 19 (38.8%). Interestingly, 26 responders had heterozygous (C/T) *Xmn1* genotype; three with homozygous (T/T); and one with wildtype (C/C). Among the non-responders, seven had heterozygous (C/T) while 12 had wildtype (C/C) *Xmn1* genotype (*p* < 0.001).

## Discussion

This study is the first to present HU response with the presence of *Xmn1* polymorphism in patients with HbE/β-thalassemia from the Bengali population of West Bengal, India. We observed that most of the transfusion-dependent patients who responded to HU therapy had heterozygous (C/T) or homozygous (T/T) *Xmn1*. Therefore, there is a significant correlation between *Xmn1* polymorphism and HU response with respect to transfusion-free interval (*p* < 0.001). Although most studies have shown almost 50% of the transfusion-dependent subjects responding to HU therapy, one of these studies revealed a high percentage (81%) of response (Kosaryan et al., 2009; Motovali-Bashi and Ghasemi, 2015; Chatterjee et al., 2018). In the present study, we found that 61.22% of the participants were good responders; this value falls within the two aforementioned percentages.

Although the present study offers major implications, it has a major limitation. We did not consider the presence of α-globin gene mutation or the roles of other genetic modifiers among the study cohort. This shortcoming can be overcome by analyzing the α-globin gene via sequencing. Therefore, our report should be considered as a pilot study until we validate the findings in a large cohort including other racial groups.

Here, we repurposed HU therapy among patients with HbE/β-thalassemia and observed a good response, especially in the patients with *Xmn1* polymorphism. We consider that the correlation between *Xmn1* polymorphism and HU therapy observed in this study is a significant predictor for possible treatment response and may have diagnostic implications.

## Data Availability

Data can be obtained on request

## Acknowledgments

The authors are grateful to the principal of Burdwan Medical College and Hospital for the continuous support. We would like to express our appreciation to the heads of the departments of Paediatrics and Pathology who facilitated this work. We thank all the patients and their families for their participation.

