## Supplementary material for "Drug repurposing: Hydroxyurea therapy improves the transfusion-free interval in HbE/beta-thalassemia–major patients with Xmn1 polymorphism": supplimentary

**Supplementary File**

**Supplementary Table 1.** Frequencies of different second beta thalassemia mutant alleles in the compound heterozygous patients with Hemoglobin E mutation[CD 26 (G>A)].

| **Sl no** | **Mutation allele 1 with**  **HGVS Nomenclature** | **Mutation allele 2 with**  **HGVS Nomenclature** | **Type of mutations for allele 2** | **Frequencies of allele 2 (%)** |
| --- | --- | --- | --- | --- |
| 1. | CD 26 (G>A)  [HBB:c.79G>A] | IVS1-5(G>C)  [HBB:c.92+5G>C] | β^0^ | 53.22 |
| 2. |  | CD30(G>C)  [HBB:c.92G>C] | β^0^ | 9.88 |
| 3. |  | CD15(G>A)  [HBB:c.47G>A] | β^0^ | 9.47 |
| 4. |  | CD15(-T)  [HBB:c.46delT] | β^0^ | 7.38 |
| 5. |  | CD41/42(-CTTT)  [HBB:c.126_129delCTTT] | β^0^ | 6.97 |
| 6. |  | IVS1-130(G>C)  [HBB:c.93-1G>C] | β^0^ | 6.55 |
| 7. |  | -90(C>T)  [HBB:c.-140C>T] | β^+^ | 6.55 |

HGVS: Human GenomeVariation Society

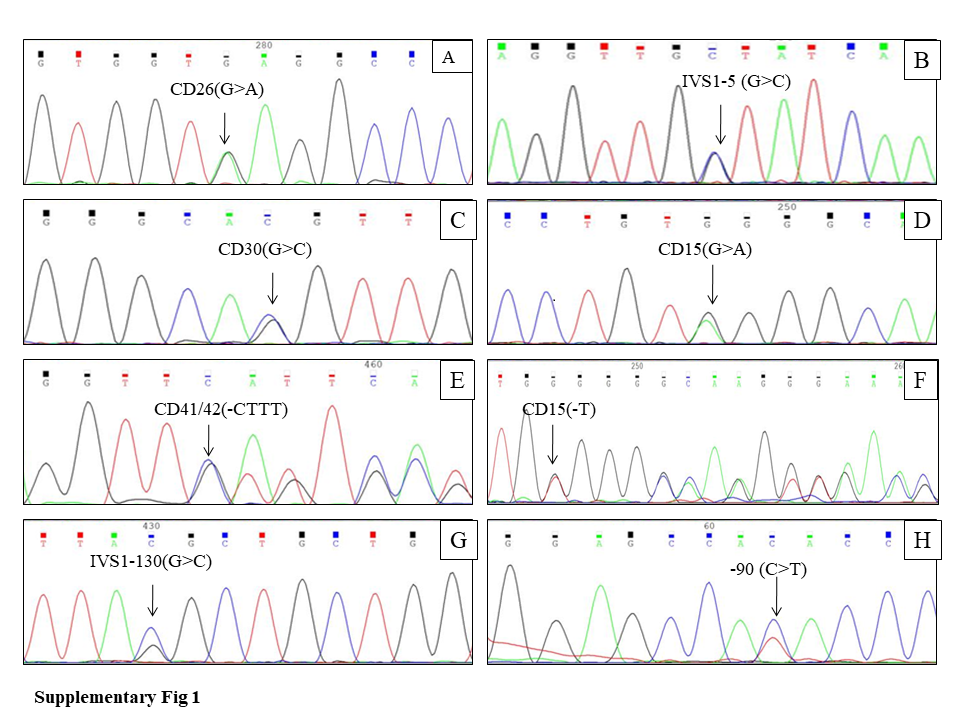

**Fig 1**. Sanger seqeuncing chromatograms showing representative (A) Mutation of CD 26(G>A) [HBB:c.79G>A], (B) IVS1-5(G>C) [HBB:c.92+5G>C], (C) CD30(G>C) [HBB:c.92G>C], (D) CD15(G>A) [HBB:c.47G>A], (E) CD41/42(-CTTT) [HBB:c.126_129delCTTT], (F) CD15(-T) [HBB:c.46delT], (G) IVS1-130(G>C) [HBB:c.93-1G>C], (H) -90 (C>T) [HBB:c.-140C>T].

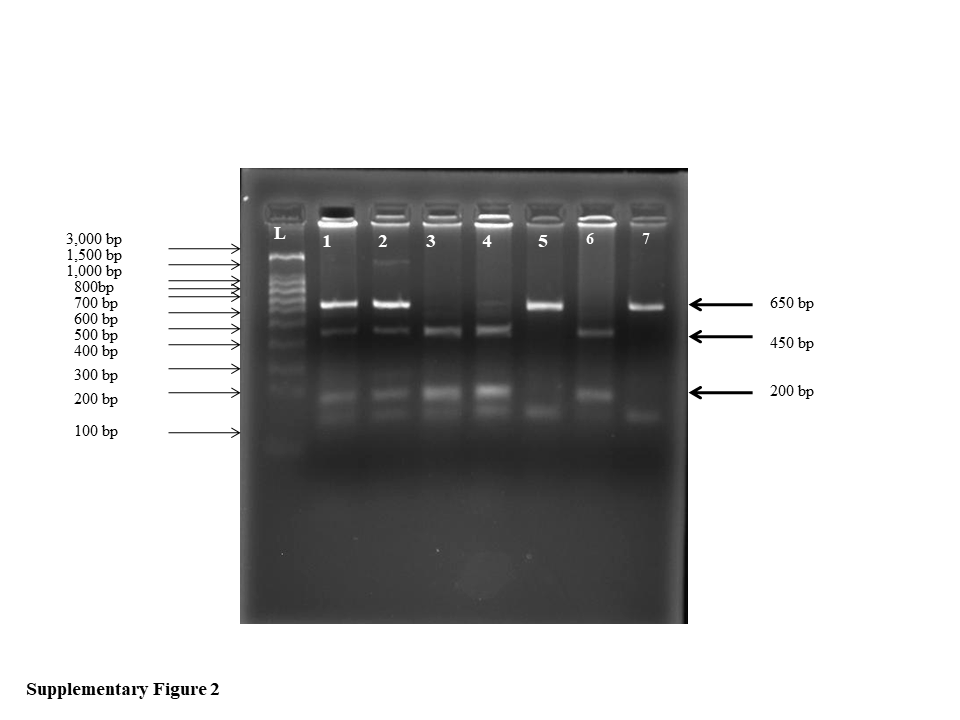

**Fig 2**. An agarose gel shows electrophoresis pattern of RFLP products of *Xmn 1 γ^G^* (-158, C>T) site. First lane (L) shows the molecular weight marker with 100 bp DNA ladder. Lane 1 and 2 show RFLP products with heterozygous genotype (C/T) for Xmn1 site (650 bp, 450 bp, 200 bp). Lane 3,4,6 show homozygous mutant genotype (T/T) (450 bp, 200 bp). Lane 5 indicates absence of *Xmn 1* site (C/C) (650 bp). Lane 7 shows undigested PCR product (650 bp).
